# Health-related hope and reduced distress associated with fluid and dietary restrictions in advanced chronic kidney disease and dialysis: a cohort study

**DOI:** 10.1101/2023.01.14.23284563

**Authors:** Noriaki Kurita, Takafumi Wakita, Shino Fujimoto, Mai Yanagi, Kenichiro Koitabashi, Masahiko Yazawa, Tomo Suzuki, Hiroo Kawarazaki, Yoshitaka Ishibashi, Yugo Shibagaki

## Abstract

**Rationale & Objective:** In chronic kidney disease (CKD), the durability of patient adherence to fluid and dietary restrictions may depend on the degree to which they have hope that they will enjoy life. However, few studies have examined the long-term role of health-related hope (HR-Hope) on distress from fluid and dietary restrictions.

Study Design Prospective cohort study.

**Setting & Participants:** A total of 444 non-dialysis stage 3-5 patients and stage 5D patients attending one of five Japanese nephrology centers.

**Exposure:** An 18-item HR-Hope scale with a score ranging from 0 to 100.

**Outcomes:** Two-item measures of restrictions on fluid and dietary intake from the Japanese version of the Kidney Disease Quality of Life Short Form, Version 1.3, with each score ranging from 0 to 100.

**Analytical Approach:** Multivariate linear mixed models were used to estimate the association of baseline HR-Hope with distress from fluid and dietary restrictions at baseline and follow-up.

**Results:** The mean age of participants was 67 years, 31.1% were female, and 124, 98, and 222 had non-dialysis CKD, peritoneal dialysis, and hemodialysis, respectively. Baseline HR-Hope buffered the growing distress from fluid restriction after one year (−18.01 pts [95% CI, -28.24 to -7.79]) per 10-pt increase, 2.59 pts [95% CI, 1.05 to 4.13]. The distress from fluid restriction at 2 years did not differ from baseline. Baseline HR-Hope buffered the growing distress from dietary restriction after one year (−12.4 pts [95% CI, -22.68 to -2.12]) per 10-pt increase, 1.96 pts (95% CI, 0.34 to 3.57). The distress from dietary restriction at 2 years did not differ from baseline.

**Limitations:** Use of self-reported measures as proxies for adherence.

**Conclusions:** Our study shows that HR-Hope, regardless of depression, can potentially mitigate the long-term distress from fluid and dietary restrictions in patients with a wide range of CKD severities.

## Introduction

Fluid and dietary restrictions constitute the cornerstone of self-management for chronic kidney disease (CKD), and their importance is emphasized by the excess risk of death due to fluid overload, hyperkalemia, and hyperphosphatemia as a result of nonadherence.^1–3^ Unfortunately, non-adherence to fluid and dietary restrictions in patients with CKD is very common, with the respective global prevalence estimates of 60.6% and 60.2% among dialysis patients.^4^ Results of several qualitative studies indicate that non-adherence to prescribed fluid and dietary restrictions is explained, in part, by a lack of motivation stemming from the impact on daily life and by psychological distress.^5–7^ Research on CKD and other chronic diseases has focused on the patients’ level of hope as an important inner source of motivation for adherence,^8–12^ however, there is insufficient evidence that hope can be a candidate for intervention in CKD.

Patient hope – which can be seen as a goal-oriented way of thinking^13,14^ – is oriented toward health^15^ and is associated with fewer negative consequences that kidney disease has on daily life, including fluid and dietary restrictions and better physiological manifestations, including blood pressure, among patients with CKD.^8,16^ However, these findings are based on cross-sectional studies and thus reverse causality cannot be excluded. Furthermore, since depression, which is prevalent among patients with CKD,^17^ has also been associated with both hope^16^ and non-adherence to fluid and dietary restrictions,^18^ the benefits of hope for fluid and dietary restrictions may be confounded by less depressive symptoms. In this context, studies examining whether health-related hope has long-term benefits for the effects of fluid and dietary restrictions independent of depression among patients with a wide range of CKD severities is clinically relevant. Specifically, such studies may help patients and clinicians to develop behavioral interventions to enhance hope and thereby promote adherence, especially when considering the paucity of evidence for long-term benefits of cognitive-behavioral therapies to increase fluid and dietary adherence.^19^

Hence, to quantify the longitudinal impact of health-related hope on the distress from fluid restriction and the distress form dietary restriction, we analyzed two-year follow-up data from a prospective cohort study – the Hope Trajectory and Disease Outcome Consortium (HOTDOC) – for Japanese patients with CKD.

## Methods

### Setting and Participants

The HOTDOC study was a multicenter cohort study conducted between February 2016 and September 2019 at five outpatient general community hospitals with nephrology services: Japanese Red Cross Medical Center (Tokyo), Inagi Municipal Hospital (Tokyo), JCHO Nihonmatsu Hospital (Fukushima), Shirakawa Kosei General Hospital (Fukushima), and St. Marianna University Hospital (Kanagawa). The HOTDOC study was conducted in accordance with the Declaration of Helsinki and was approved by the institutional review boards of Fukushima Medical University (number 2417) and St. Marianna University (number 3209). Written informed consent was obtained from each participant. Inclusion criteria were as follows: (1) stage 5D CKD receiving hemodialysis or peritoneal dialysis therapy at the participating centers, or (2) non-dialysis stage 3-5 CKD receiving nephrology care at the participating centers, for dietary instruction, medication prescription, and/or kidney function monitoring, and (3) ability to complete the questionnaire survey. Patients with dementia were excluded.

### Exposure

The main exposure is hope, as measured by the Health-Related Hope (HR-Hope) scale, which assesses the hope related to health among people with chronic illness.^15^ This scale consists of 18 items and is unidimensional. Three subdomains can be scored via structural validation: “something to live for” (5 items), “health and illness” (6 items), and “role and connectedness” (7 items). Responses to each item were scored on a four-point Likert scale with scores ranging from one (“I don’t feel that way at all”) to four (“I strongly feel that way”). The average scores for the total domain and each subdomain were obtained and converted to a scale ranging from 0 to 100 points. Patients without a family member were waived from answering two items (both in the “roles and connectedness” subdomain). The scale had been demonstrated to have adequate reliability (Cronbach’s alpha coefficient: 0.93), as well as criterion validity and construct validity.^15^

### Outcomes

The main outcomes were distress from restriction on fluid and distress from restriction on dietary intake as proxies for nonadherence to fluid restriction and dietary restriction, respectively. These two items are included in an eight-item subscale of Effects of Kidney Disease on Daily Life, as measured by the Japanese version of the Kidney Disease Quality of Life Short Form (KDQOL-SF™), Version 1.3.^20^

The items “Fluid restriction?” and “Dietary restriction?” were presented after the following instructional text: “How much does kidney disease bother you in each of the following areas?” The patients chose one response from a scale of 1 (Not at All bothered) to 5 (Extremely bothered).

The chosen raw scores for the items were inverted to a range of 0-100, with higher scores reflecting a higher quality of life.^21^ The two items were treated as separate scores in the present study.

These items were collected at baseline, after one year, and after two years.

### Covariates

Age, sex, diabetic nephropathy, stage of kidney disease and treatment status (non-dialysis, peritoneal dialysis, hemodialysis), comorbidities (coronary artery disease, cerebrovascular disease, and malignancy), serum potassium, serum phosphorus, systolic blood pressure, potassium binder, number of phosphorus binders, and number of anti-hypertensive classes were collected from medical records.

The stage of CKD was determined by estimated glomerular filtration rate. Dialysis vintage was also collected. Performance status was assessed using the Zubrod scale by attending physicians.^22^ Poor performance status was defined as a score of 2 (walking more than 50% of the awake time or receiving occasional assistance when moving) or higher. The presence of family was evaluated by the patient’s yes or no response to the question “Do you have any family?” in the HR-Hope scale.^15^ Working status was used as a proxy for economic status and measured using the item “During the past 4 weeks, did you work at a paying job?” in the KDQOL to which patients responded yes or no.^20^ Depression was assessed using the Japanese version of the Center for Epidemiologic Studies Depression scale (CES-D).^23,24^ The CES-D consists of 20 items and can be scored using a 4-point scale. The total score ranges from 0 to 60, and a total score of 16 or higher was considered depressed.^17,23^

The questionnaire was administered at each facility, and patients were asked to complete it at home or during their nephrology visit. In the event that patients could not write due to visual impairment or physical disability, they were asked to verbally complete the form with the aid of a trained research assistant who did not inform patients of the hypothesis.

### Statistical Analysis

All statistical analyses were performed in Stata/SE version 17. For baseline descriptive statistics, continuous variables were summarized by mean and standard deviation or median and interquartile range, and categorical variables were summarized by frequency and percentage. They were presented for the total population and by treatment status. A Sankey diagram was created to provide a graphical overview of the distribution of values for water and dietary restriction scores and the frequency of their combination.^25^

Linear mixed models were fitted to examine the association between baseline outcome scores and HR-Hope and whether changes in those scores over time differed by HR-Hope score. The models were fitted for each fluid restriction and diet restriction item. Robust variance estimation was used to address heteroscedasticity. The covariates described above were forced into the models. Serum potassium, serum phosphorus, potassium binder, and the number of phosphorus binders were included only in the model for dietary restriction, while the systolic blood pressure and the number of antihypertensive medication classes were included only in the model for fluid restriction. Stage of kidney disease and renal replacement therapy status were combined into one variable, consisting of six CKD levels: stage 3, stage 4 or 5, peritoneal dialysis within 1 year, peritoneal dialysis >1 year, hemodialysis within 1 year, and hemodialysis >1 year. The Wald test was used to examine whether the temporal changes in the outcome scores differed by HR-Hope score. Predicted means for the outcome scores were calculated to depict temporal changes in the scores by HR-Hope score. The scores represented those as if the total population had the same time points and HR-Hope scores, while the distribution of covariates for the total population remained unchanged.^26^

For any covariate with missing values, multiple imputation with chained equations was performed assuming that the missing values were at random.^27^ Estimates from 10 imputed data were combined into a single estimate.

## Results

### Baseline characteristics

After excluding 17 cases with missing values for either baseline HR-Hope (n = 3) or fluid restriction/dietary restriction (n = 15), 444 participants were included in the analysis. Baseline characteristics of the participants are shown in Table 1. Among 124 non-dialysis, 53, 52, and 19 had stage 3, stage 4 or 5, respectively. Among 98 peritoneal dialysis, 19 and 79 were within 1 year and >1 year, respectively. Among 222 hemodialysis, 47 and 175 were within 1 year, and hemodialysis >1 year, respectively. A total of 10.4% of participants had decreased performance status and 32.7% had depressive states.

**Table 1.** Baseline characteristics of the study participants (n = 444)

|  | Treatment categories |  |  | Total |
| --- | --- | --- | --- | --- |
|  | *Non-dialyzed* | *Peritoneal dialysis* | *Hemodialysis* |  |
|  | n = 124 | n = 98 | n = 222 | n = 444 |
| **<u>Demographics</u>** |  |  |  |  |
| **Age, years<sup>b</sup>** | 72.7 (12.4) | 66.5 (13.5) | 64 (14) | 67 (13.9) |
| **Female** | 39 (31.5 %) | 28 (28.6 %) | 71 (32 %) | 138 (31.1 %) |
| **Vintage, month<sup>b</sup>** | n.a. | 33.4 [13.7 - 64.3] | 54.9 [16.2 - 120.1] | 45.8 [15.7 - 98.2] |
| **Renal disease** |  |  |  |  |
| *Diabetic nephropathy* | 16 (12.9 %) | 25 (25.5 %) | 75 (33.8 %) | 116 (26.1 %) |
| *Glomerulonephritis* | 18 (14.5 %) | 37 (37.8 %) | 60 (27 %) | 115 (25.9 %) |
| *Hypertensive disease* | 41 (33.1 %) | 9 (9.2 %) | 32 (14.4 %) | 82 (18.5 %) |
| *Others* | 49 (39.5 %) | 27 (27.6 %) | 55 (24.8 %) | 131 (29.5 %) |
| **Having family, yes** | 112 (91.8 %) | 83 (84.7 %) | 201 (90.5 %) | 396 (89.6 %) |
| *Missing, n* | 2 | 0 | 0 | 2 |
| **Working, yes** | 83 (67.5 %) | 56 (57.1 %) | 157 (70.7 %) | 296 (66.8 %) |
| *Missing, n* | 1 | 0 | 0 | 1 |
| **Impaired performance status, yes** | 5 (4 %) | 8 (8.2 %) | 33 (14.9 %) | 46 (10.4 %) |
| **Depression** | 23 (20.4 %) | 37 (38.5 %) | 80 (36.5 %) | 140 (32.7 %) |
| *Missing, n* | 11 | 2 | 3 | 16 |
| **<u>Comorbidities</u>** |  |  |  |  |
| **Coronary artery disease** | 17 (13.7 %) | 19 (19.4 %) | 32 (14.4 %) | 68 (15.3 %) |
| **Cerebrovascular disease** | 16 (12.9 %) | 16 (16.3 %) | 28 (12.6 %) | 60 (13.5 %) |
| **Malignancy** | 13 (10.5 %) | 9 (9.2 %) | 24 (10.8 %) | 46 (10.4 %) |
| **SBP, mmHg** | 133.2 (18) | 132.3 (22.6) | 148.4 (25.9) | 141.1 (24.7) |
| *missing, n* | 26 | 1 | 0 | 27 |
| **Potassium, mEq/L** | 4.6 (0.5) | 4.2 (0.6) | 4.8 (0.7) | 4.6 (0.7) |
| *missing, n* | 3 | 0 | 1 | 4 |
| **Phosphorus, mg/dL** | 3.6 (0.6) | 5.3 (1.4) | 5.5 (1.4) | 5 (1.5) |
| *missing, n* | 31 | 0 | 1 | 32 |
| **Prescription of potassium binder, %** | 9 (7.3 %) | 3 (3.1 %) | 52 (23.4 %) | 64 (14.4 %) |
| **N of phosphate binder, %** |  |  |  |  |
| *None* | 122 (98.4 %) | 17 (17.4 %) | 80 (36 %) | 219 (49.3 %) |
| *1 to 3* | 1 (0.8 %) | 11 (11.2 %) | 45 (20.3 %) | 57 (12.8 %) |
| *4 to 6* | 1 (0.8 %) | 28 (28.6 %) | 43 (19.4 %) | 72 (16.2 %) |
| *7 or over* | 0 (0 %) | 42 (42.9 %) | 54 (24.3 %) | 96 (21.6 %) |
| **N of categories for antihypertensives, %** |  |  |  |  |
| *None* | 33 (26.6 %) | 37 (37.8 %) | 65 (29.3 %) | 135 (30.4 %) |
| *1* | 36 (29 %) | 29 (29.6 %) | 51 (23 %) | 116 (26.1 %) |
| *2 or over* | 55 (44.4 %) | 32 (32.7 %) | 106 (47.8 %) | 193 (43.5 %) |
| **HR-Hope, points<sup>a, b</sup>** | 65.3 (16.5) | 62.9 (17.4) | 57.8 (19.4) | 61 (18.5) |
| **Effect of water restriction, points<sup>b</sup>** | 97.6 (7.4) | 78.8 (25) | 63.4 (32.2) | 76.4 (29.7) |
| **Effect of diet restriction, points<sup>b</sup>** | 79.8 (25) | 70.4 (26) | 73.1 (29) | 74.4 (27.5) |

<sup>a</sup>Health-related hope score (i.e., lower score indicates worse hope related to health).
<sup>b</sup>Values for continuous data are shown as mean (standard deviation) and/or median [25th and 75th percentiles].

Compared to non-dialysis patients, dialysis patients were younger, were more likely to have diabetic nephropathy, had lower performance status, had higher systolic blood pressure and serum phosphorus, and a greater proportion were depressed. Compared to non-dialysis patients, more dialysis patients were prescribed potassium and phosphorus binders.

The mean HR-Hope score, fluid restriction, and dietary restriction scores were 61, 76.4, and 74.4, respectively. Hemodialysis patients had the worst HR-Hope and fluid restriction scores.

The distribution of values for baseline fluid and dietary restriction scores and frequency of their combination across the whole population is shown in a Sankey diagram (Figure 1). Most patients rated the fluid and dietary restriction equally, while some rated the fluid restriction as not bothersome but the dietary restriction as bothersome, and Vice versa.

**Figure 1.**
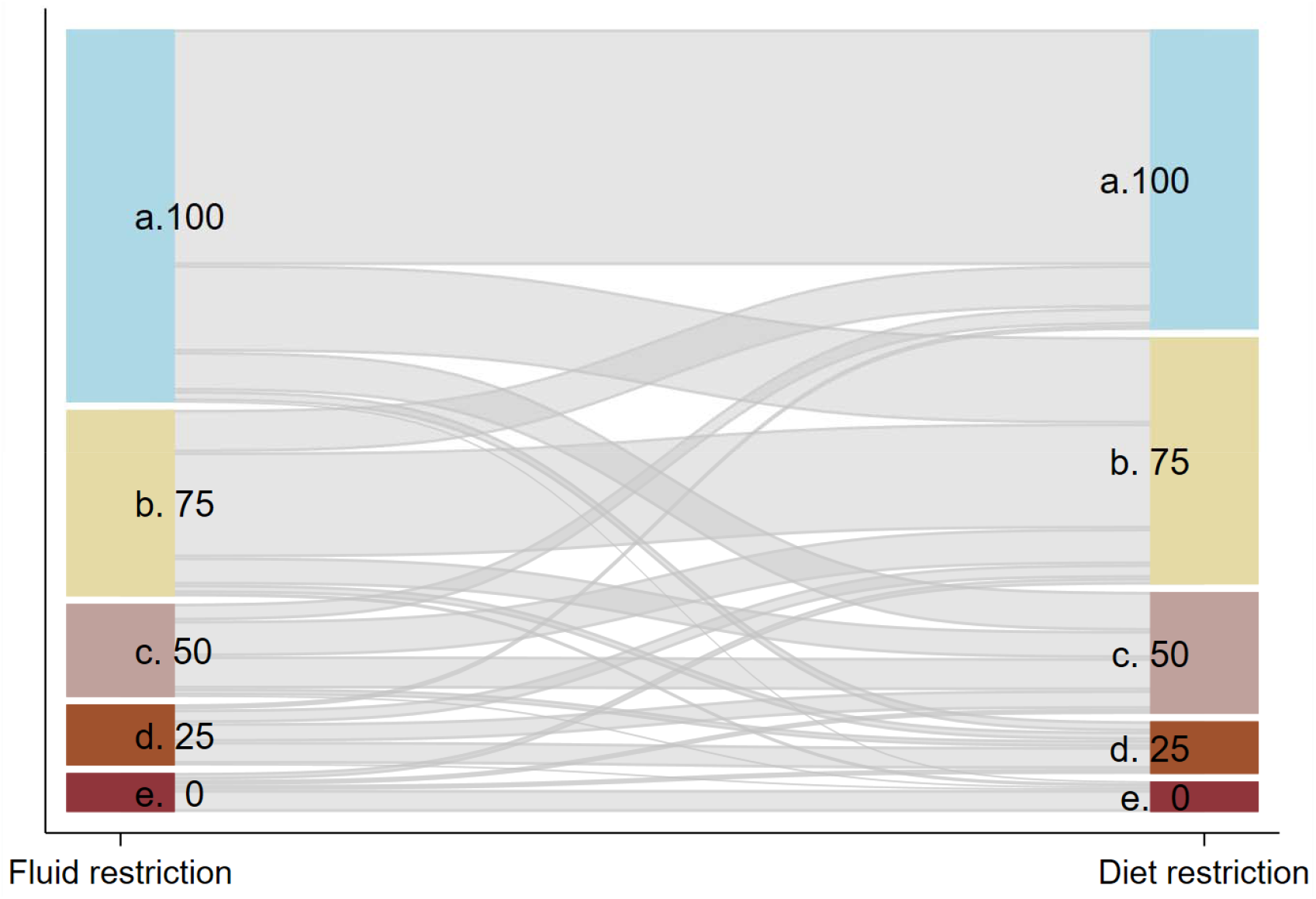
Distribution and combination of fluid and dietary restriction scores at baseline presented by a Sankey diagram. A Sankey diagram was used to depict the combination (flow) of the values of the fluid and dietary restriction scores measured by the items from the Kidney Disease Quality of Life Short Form (KDQOL-SF™), Version 1.3. The height of individual boxes (nodes) on the vertical axis indicates relative proportions. The thickness of the links connecting the boxes for the effects of fluid and dietary restrictions indicates the relative proportions of the combination. Light blue indicates “Not at All bothered” (100 points), light khaki indicates “Somewhat bothered” (75 points), rose indicates “Moderately bothered” (50 points), sienta indicates “Very Much bothered” (25 points), and maroon indicates “Extremely bothered” (0 points).

### Follow up

A total of 149 patients did not participate in the 1-year follow-up survey because of referral to other facilities (n = 31), death (n = 25), hospitalization (n = 4), change in treatment modality (n = 1), or unknown reasons (n = 88). In addition, 241 patients did not participate in the 2-year follow-up survey because of death (n = 48), referral to other facilities (n = 42), hospitalization (n = 4), change in treatment modality (n = 1), or unknown reasons (n = 146). Thus, 295 and 203 patients were included in the longitudinal analysis at 1 and 2 years, respectively.

### Associations of fluid restriction score with HR-Hope, timeline, and covariates

Findings demonstrating how baseline HR-Hope, timeline, and covariates were associated with the fluid restriction score are presented in Table 2.

**Table 2.** Associations of water restriction score with hope and covariates† (n = 444, observations =941)

|  | Corresponding standardised ES | Mean difference, point estimate (95%CI) | P-value |
| --- | --- | --- | --- |
| Health-related hope, per 10-pt increase | 0.06 | 1.08 (-0.3 to 2.46) | 0.125 |
| Timeline |  |  |  |
| Baseline |  | Reference |  |
| 1-yr after | **-0.98** | **-18.01 (-28.24 to -7.79)** | **0.001** |
| 2-yr after | 0.36 | 6.7 (-6.21 to 19.61) | 0.309 |
| Interactions<sup>a</sup> |  |  |  |
| 1-yr after × Health-related hope, per 10-pt increase | **0.14** | **2.59 (1.05 to 4.13)** | **0.001** |
| 2-yr after × Health-related hope, per 10-pt increase | -0.07 | -1.29 (-3.33 to 0.74) | 0.212 |
| Age, per 10-yr increase | **0.14** | **2.51 (0.98 to 4.04)** | **0.001** |
| Female vs. Male | 0.06 | 1.13 (-3.35 to 5.61) | 0.621 |
| Diabetic Nephropathy | -0.04 | -0.83 (-5.88 to 4.23) | 0.749 |
| Stage of kidney disease |  |  |  |
| Non-dialysis stage 2/3 |  | Reference |  |
| Non-dialysis stage 4/5 | 0.07 | 1.35 (-2.6 to 5.3) | 0.503 |
| PD, >0 to 1 yr | **-0.99** | **-18.23 (-29.6 to -6.86)** | **0.002** |
| PD, >1 yr | **-0.76** | **-13.95 (-18.89 to -9.02)** | **<0.001** |
| HD, >0 to 1 yr | **-1.67** | **-30.75 (-37.72 to -23.79)** | **<0.001** |
| HD, >1 yr | **-1.57** | **-28.93 (-33.84 to -24.02)** | **<0.001** |
| Impaired performance status | 0.38 | 7.09 (-1.01 to 15.2) | 0.086 |
| Having family | 0 | -0.01 (-4.87 to 4.86) | 0.998 |
| Working | 0.18 | 3.29 (-1.24 to 7.82) | 0.155 |
| Coronary artery disease | 0.03 | 0.5 (-5.81 to 6.81) | 0.877 |
| Cerebrovascular disease | -0.35 | -6.49 (-13.4 to 0.42) | 0.066 |
| Malignancy | 0.13 | 2.32 (-4.1 to 8.73) | 0.480 |
| Depressive symptom | **-0.45** | **-8.27 (-13.09 to -3.45)** | **0.001** |
| SBP, per 10-mmHg increase | 0.01 | 0.17 (-0.76 to 1.1) | 0.719 |
| N of categories for antihypertensives |  |  |  |
| None |  | Reference |  |
| 1 | -0.16 | -2.93 (-7.78 to 1.92) | 0.236 |
| 2 or over | **-0.31** | **-5.73 (-10.65 to -0.82)** | **0.022** |

<sup>†</sup>Linear mixed effect model with robust variance estimation was fit with inclusion of all variables listed above.
<sup>a</sup>Each interaction term models the impact of the baseline health-related hope score on the restriction score at each timeline.
PD: Peritoneal dialysis, HD: Hemodialysis, SBP: Systolic blood pressure

HR-Hope modified temporal changes in the fluid restriction score (*P* < 0.001 for interaction by Wald test). Baseline HR-Hope was not associated with the fluid restriction score at baseline (per 10-pt increase, 1.08 pts; 95% confidence interval [95% CI] -0.3 to 2.46). The fluid restriction score at one year deteriorated (−18.01 pts, 95% CI -28.24 to -7.79). However, baseline HR-Hope mitigated the deteriorating fluid restriction score at one year (per 10-pt increase, 2.59 pts, 95% CI 1.05 to 4.13). The fluid restriction score at 2 years did not differ from baseline (6.7 pts, 95% CI -6.21 to 19.61).

Older age (per 10-year incremental increase) was associated with a higher baseline fluid restriction score (2.51 pts, 95% CI 0.98 to 4.04). Depression and prescription of two or more categories of anti-hypertensive drugs were associated with a lower fluid restriction score at baseline (−8.27 pts, 95% CI -13.09 to -3.45; -5.73 pts, 95% CI -10.65 to -0.82, respectively). Compared to non-dialysis stage 2/3, peritoneal dialysis and hemodialysis had a lower fluid restriction score at baseline.

The predicted mean scores for fluid restriction at specific HR-Hope scores at baseline, derived from the linear mixed-effects models in Tables 2, are shown in Figure 2A. At a baseline HR-Hope score of 0, the fluid restriction score at 1 year was deteriorated (50.5 pts, 95% CI 39.8 to 61.1). At a baseline HR-Hope score of 80, no deterioration in the fluid restriction score was observed at 1 year (79.8 pts, 95% CI 76.1 to 83.5).

**Figure 2.**
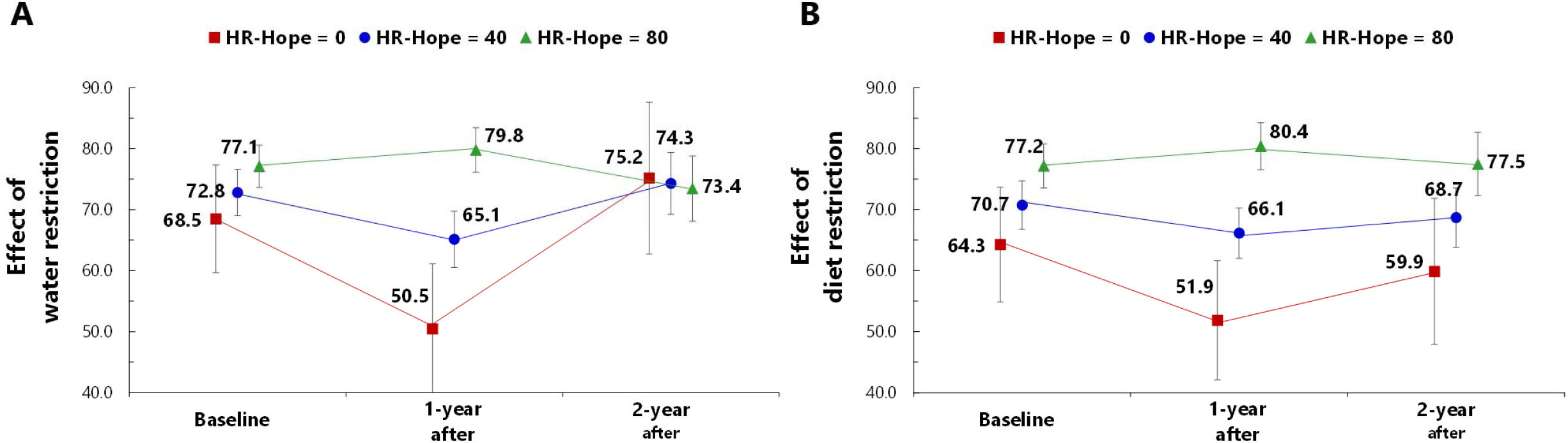
Evolution of predicted mean scores for fluid and diet restriction scores by selected baseline hope scores. Predicted mean values of the fluid (A) and dietary (B) restriction scores at selected HR-Hope scores at baseline were derived from the linear mixed effect models presented in Table 2 and Table 3, respectively. Green triangles, blue circles, and red squares indicate point estimets of the effects scoers for the patients having baseline HR-Hope scores of 0, 40, and 80 points, respectively. Error bars indicate 95% confidence intervals. HR-Hope: health-related hope

### Associations of dietary restriction score with HR-Hope, timeline, and covariates

Findings on how baseline HR-Hope, timelne, and covariates were associated with the dietary restriction score are presented in Table 3.

**Table 3.**
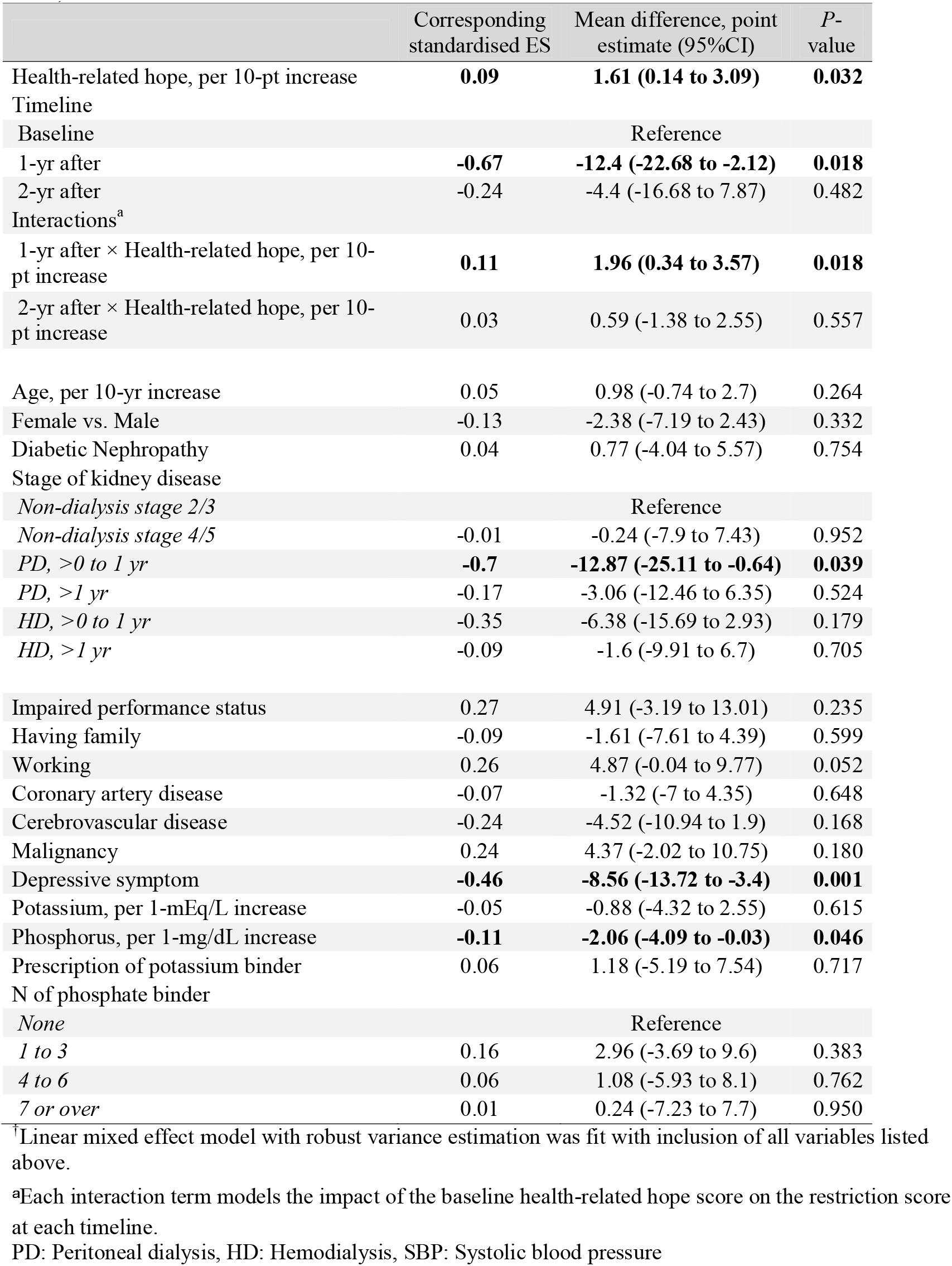
Associations of diet restriction score with hope and covariates† (n = 444, observations =942)

HR-Hope modified temporal changes in the dietary restriction score (*P* = 0.05 for interaction by Wald test). Higher baseline HR-Hope was associated with a higher dietary restriction score at baseline (per 10-pt increase, 1.61 pts, 95% CI 0.14 to 3.09). While the dietary restriction score at one year deteriorated (−12.4 pts, 95% CI -22.68 to -2.12), baseline HR-Hope mitigated the deteriorating dietary restriction score at one year (per 10-pt increase, 1.96 pts, 95% CI 0.34 to 3.57). The dietary restriction score at 2 years did not differ from baseline (−4.4 pts, 95% CI -16.68 to 7.87).

Depression and serum phosphorus were associated with lower dietary restriction score at baseline (−8.56 pts, 95% CI -13.72 to -3.4; per 1 mg/dl increase, -2.06 pts, 95% CI -4.09 to -0.03, respectively). Compared to non-dialysis stage 2/3, peritoneal dialysis within 1 year had a lower dietary restriction score at baseline (−12.87 pts, 95% CI -25.11 to -0.64).

The predicted mean scores for the dietary restriction at specific HR-Hope scores at baseline, derived from the linear mixed-effects models in Tables 3, are shown in Figure 2B. At a baseline HR-Hope score of 0, the dietary restriction scores at 1 year were deteriorated (51.9 pts, 95% CI 42.1 to 61.6). At a baseline HR-Hope score of 80, no deterioration in the dietary restriction score was observed at 1 year (80.4 pts, 95% CI 76.5 to 84.3).

## Discussion

In the present study, we investigated whether health-related hope plays a role in the effects of fluid and dietary restrictions on day-to-day life among patients with CKD. Our results showed that higher health-related hope mitigated the worsening of both the fluid and dietary restriction scores after 1 year, independent of depressive symptoms.

Our first finding that health-related hope of patients with CKD appears to play a role in the effects associated with fluid and dietary restrictions is supported by our two-year longitudinal study: the increments of higher baseline health-related hope requiring a moderate standardized effect size to buffer worsening after one year were 30 (0.14 × 3 = 0.42) and 40 (0.11 × 4 = 0.44) points for fluid and dietary restriction, respectively.^28^ The finding extends those of previous observational studies of hope in patients with CKD. First, both the study showing high general hope and low impact of kidney disease on daily life in dialysis patients and the study showing high health-related hope and low impact of fluid and dietary restrictions in a wide range of CKD severities were based on cross-sectional studies.^8,16^ The present finding showing the longitudinal relationship is unlikely to demonstrate reverse causality, in which the high impact on daily life results in hopelessness. However, a relationship of mitigating the deterioration of the restriction score by health-related hope was not observed at two years. Whether this is due to patient accommodation to the restrictions is unclear.

Our second finding is that the possibility of mitigating the fluid and dietary restrictions scores by hope is independent of depression: this has not been addressed in previous studies. Given that low hope is clearly associated with high depressive states^15,16^ and that depression is associated with poor adherence to fluid and dietary restrictions,^18^ previous studies of hope and low impact of fluid and dietary restrictions may have been confounded by less depressive states.

The findings of this study have implications for researchers and clinicians on several aspects. First, the fluid and dietary restrictions scores are potentially modifiable factors and could be reduced by enhancing hope through psychological interventions such as cognitive-behavioral therapy. Systematic reviews have shown that cognitive-behavioral therapy to increase fluid and dietary adherence may be effective, but the effects persist at best for up to a year.^19^ Of these interventions, one that demonstrated the most long-lasting effects was a simple program consisting of self-affirming manipulations such as recalling past acts of kindness and providing health risk information regarding renal care, resulting in reduced serum phosphorus or weight gain, but there were no improvements in self-efficacy or behavioral intentions.^29,30^ Based on our study, we can propose cognitive behavioral therapy that incorporates the identification of health-related hope inherent in patients with CKD as well as the specific healthy behaviors required to achieve it, formulating plans to implement those behaviors, and counseling on their barriers and concerns. Indeed, one study showed an increase in hope in hemodialysis patients through group counseling that reduced existential distress and corrected cognitive errors that leave undesirable behaviors unchanged.^31^ Another study showed reductions in stress in hemodialysis patients that was achieved through structured, repetitive counseling to help them discover what their hopes were and find pathways and remove barriers to achieving them.^32^ Further research is warranted to establish whether long-term maintenance of hope and potential mitigation of the distress from fluid and dietary restrictions can demonstrate improvements in self-efficacy, healthy behaviors, and objective adherence measures.

Second, this study raises the need for clinicians to inquire about whether health-related hope is compromised even in the absence of depression during their routine dialogue with patients with kidney disease. While hopelessness is considered part of depression, hopelessness can be quite common in the absence of depression,^33,34^ and also hopelessness may not be recognized as a symptom of depression but rather be considered only as a risk factor for depression.^35^ Third, the finding that other measures of fluid and dietary adherence are related to each restriction score may contribute to a revision for patient instruction. The finding that being prescribed two or more antihypertensive drug categories has a small standardized effect size on the fluid restriction score suggests that patients may have conflicts in their daily lives linked to fluid intake and treatment decisions with their healthcare provider when managing blood pressure. In addition, since serum phosphorus exceeding well above the upper limit of the management goal may have a small standardized effect size on dietary restriction score (e.g., for each 2 mg/dl increase, -0.11 × 2 = -0.22),^28^ frequent laboratory feedback may instead be potentially threatening to patients who are unable to adhere to recommended dietary restrictions.^30^ This contradicts the fact that patients who are confident in their dietary strategy are highly motivated to adjust their diet in response to laboratory test results.^5^ Thus, even if frequent laboratory tests could help achieve guideline targets,^36^ it may be necessary to consider the patient’s individuality in order to provide effective feedback.

The strength of our study is that we were able to examine the 2-year long-term impact of hope on the distress from fluid and dietary restrictions in a wide range of CKD severity, with an adjustment for depression. In addition, the multicenter study design ensures the generalizability of our findings.

Several limitations of this study warrant a mention. First, the outcome of this study was solely based on the self-reported distress from fluid and dietary restrictions as a surrogate indicator of adherence. We could not measure fluid intake by volume nor dietary-derived phosphorus intake. However, it should be noted that biochemical and physiological surrogate measures such as phosphate and interdialytic weight gain are also widely used as indicators of adherence in dialysis patients.^19^ Second, a modest percentage of patients were lost to follow-up due to referral to other facilities, death, or other unknown reasons, and thus their response to distress from fluid and dietary restrictions were missed during the follow-up period.

In summary, our study shows that health-related hope can potentially mitigate the long-term distress from fluid and dietary restrictions in patients with a wide range of CKD severities. Since hope is an easily overlooked aspect of patient psychology in the context of practice guidelines centered on laboratory testing and physiologic indicators, we need to recognize hope as a target that must be considered when developing new strategies to improve adherence.

## Author Contribution

Research idea and study design: NK, TW, YI, YS; data acquisition: NK, SF, M. Yanagi, TS, KK, M. Yazawa, HK, YI, YS; data analysis/interpretation: NK, TW, TS, M. Yazawa, HK, YI, YS; statistical analysis: NK; supervision: TW, YI, YS.

Each author contributed important intellectual content during manuscript drafting or revision and agrees to be personally accountable for the individual’s own contributions and to ensure that questions pertaining to the accuracy or integrity of any portion of the work, even one in which the author was not directly involved, are appropriately investigated and resolved, including with documentation in the literature if appropriate.

## Data Availability

All data produced in the present work are contained in the manuscript.

## Acknowledgement

The authors greatly thank the following researchers, research assistants, and medical staff members for their assistance in collecting the questionnaire-based and clinical information used in this study: Ms. Asako Tamura, Ms. Yuka Masuda, and Ms. Takae Shimizu (St. Marriana University, Kawasaki-City, Kanagawa); Takayuki Nakamura, MD and Eiko Hashimoto, RN (JCHO Nihonmatsu Hospital, Nihonmatsu-City, Fukushima); Atsushi Kyan, MD and Masashi Saito, CE (Shirakawa Kosei General Hospital, Shirakawa-City, Fukushima); Ms. Lisa Shimokawa and Ms. Miyuki Sato (Fukushima Medical University Hospital, Fukushima-City, Fukushima).

